# Isolevuglandins promote autoimmunity and hypertension in systemic lupus erythematosus

**DOI:** 10.1101/2020.02.10.20021741

**Authors:** David M. Patrick, Nestor de la Visitación, Michelle J. Ormseth, C. Michael Stein, Sean S. Davies, Valery N. Yermalitksy, Venkataraman Amarnath, Leslie J. Crofford, Jonathan M. Williams, Sergey Dikalov, Anna Dikalova, Liang Xiao, Justin P. Van Beusecum, Mingfang Ao, Agnes B. Fogo, Annet Kirabo, David G. Harrison

## Abstract

Hypertension, vascular inflammation and renal inflammation are characteristic of systemic lupus erythematosus (SLE), a multisystem autoimmune disease that is complex and poorly understood. Oxidation products of arachidonic and other fatty acids, termed isolevuglandins (isoLG) lead to formation of post-translational protein modifications that are immunogenic. We demonstrate isoLG enrichment in dendritic cells (DCs), B cells, and plasma cells from juvenile female *B6.SLE123* mice. In adult *B6.SLE123* and *NZBWF1* mice, isoLG adducts are enriched in plasma cells and splenic DCs compared to *C57Bl/6* and *NZW* mice respectively. Treatment with the isoLG-scavenger 2-hydroxybenzylamine (2-HOBA) reduced blood pressure, improved renal function, and attenuated renal injury. Moreover, 2-HOBA reduced bone marrow plasma cells, total IgG levels, and anti-dsDNA antibody titers. We also demonstrate that mice with SLE generate specific IgG antibodies against isoLG adducted protein, confirming the immunogenicity of isoLG adducts. Finally, we found that isoLG adducted peptides are markedly enriched in monocytes from patients with SLE which was accompanied by an increase in superoxide production. These findings support a role of isoLG adducts in the genesis and maintenance of systemic autoimmunity and its associated hypertension in SLE. Scavenging of isoLGs promises to be a novel therapy for this disease.

## INTRODUCTION

Systemic lupus erythematosus (SLE) is a multi-organ autoimmune disease that affects approximately nine females for every male (1). Cardiovascular disease, particularly myocardial infarction, stroke, and hypertension are increased in SLE (2). In women with SLE the prevalence of hypertension is higher when compared to healthy controls (3). The mechanism of hypertension in SLE has been attributed to multiple factors including concomitant renal disease and side effects of medications but remains unexplained. Importantly, in a study of 235 patients with SLE, hypertension was not correlated with renal disease or serum complement levels, suggesting that the occurrence of hypertension in SLE can be independent of renal disease (4).

Aside from the hypertension related to autoimmune disease, immune activation has also been implicated in both humans with essential hypertension and in several models of experimental hypertension (5). Immune cells, including macrophages, T cells, and B cells are present in the kidneys and vasculature. Cytokines released by these cells are implicated in vascular remodeling, vasoconstriction and sodium retention (5-8). Mice that lack T cells have a blunted hypertensive response to angiotensin II (ang II) or deoxycorticosterone acetate, and deletion of the RAG1 gene blunts hypertension and end-organ damage in rats with salt-sensitive hypertension (9). Furthermore, blockade of T cell costimulation by CTLA4-Ig inhibited the development of hypertension in mice, suggesting a critical role of antigen presenting cells (APCs), such as monocytes and DCs, in the initiation of hypertension (10).

Interestingly, excessive levels of reactive oxygen species (ROS) have been implicated in both hypertension and SLE. Oxidation-induced apoptosis has been linked to formation of autoantigens in SLE, and autoantibody production is stimulated by oxidative stress (11-14). ROS generation in SLE has been attributed to non-synonymous polymorphisms in genes encoding mitochondrial complex I and V (15). ROS contribute to essential hypertension via renal, vascular, and central mechanisms(16). In the *NZWBWF1* mouse model of SLE, treatment at 30 weeks of age with the antioxidant tempol and the NADPH oxidase inhibitor apocynin reduced blood pressure and albuminuria. Such treatment reduces urinary and renal cortical hydrogen peroxide, suggesting that a reduction in oxidative stress attenuates hypertension in SLE (17).

Recently, we and others have elucidated a new mechanism in the pathogenesis of hypertension by ROS. Hypertension is associated with modification of self-proteins by isolevuglandins (isoLGs), which are *γ*-ketoaldehydes derived from the oxidation of fatty acids and phospholipids (18). IsoLGs react with free primary amines leading to covalent modification of lysine residues on proteins. IsoLG adducts may be detected by flow cytometry utilizing the single chain antibody D11 ScFv that recognizes isoLG-lysine adducts on any protein (19). In several mouse models of hypertension, isoLG adducts accumulate within APCs and promote their maturation and activation (20). Small molecule scavengers of isoLGs, including 2-hydroxybenzylamine (2-HOBA), lower blood pressure and markedly reduce end-organ damage in both ang II and DOCA-salt hypertension (20). It is conceivable that isoLG adduction of peptides plays a similar role in SLE, however, this has not been investigated. In this regard, serum from the lupus prone *MRL/lpr* mouse reacts with bovine serum albumin (BSA) that has been similarly adducted by the highly reactive *γ*-ketoaldehyde 4-oxo-2-nonenol (ONE) and ONE-modified BSA is recognized by anti-DNA antibodies prepared from serum of lupus prone mice (14). In the present study, we examined monocytes from humans and employed two mouse models of SLE to define a role of isoLG formation in the pathogenesis of hypertension and systemic autoimmunity in SLE.

## RESULTS

### *B6.SLE123* mice exhibit augmented isoLG adduct accumulation in addition to derangements in immune cell populations

We hypothesized that isoLG adduct accumulation occurs before the onset of overt systemic autoimmunity. The progression of disease in *B6.SLE123* animals has been previously characterized, and importantly, these animals do not display an increase in mortality before 4-months of age (21). We therefore chose to study 7-week old *B6.SLE123* animals by flow cytometry using gating strategies outlined in the supplemental figures (Supplemental Figures 1-3). We found an increased number of plasma cells in the bone marrow and that isoLG adducts are enriched in these cells (Figure 1 A-D). We also observed an accumulation of DCs in the spleen in addition to isoLG adduct accumulation within splenic DCs (Figure 1 E-G). We also observed plasma cell accumulation in spleen similar to that of bone marrow (Figure 1 H-I). Importantly, splenic accumulation of total CD45^+^ cells and CD3^+^ T cells are unchanged at this age (Figure 1 J-K). Finally, we observed that while *B6.SLE123* mice had fewer splenic B cells, these cells also displayed marked accumulation of isoLG adducts (Figure 1 L-N). We did not observe renal immune cell infiltration at this age (Figure 1 O-P). In adult 32 week old *B6.SLE123* animals, we discovered that isoLG adducts accumulate in peripheral activated B cells when compared to naïve B cells (Figure 2 A-C). CD44 is an adhesion molecule present on numerous leukocytes. On B cells, CD44 is upregulated following antigen priming and activation (22, 23).

**Figure 1:**
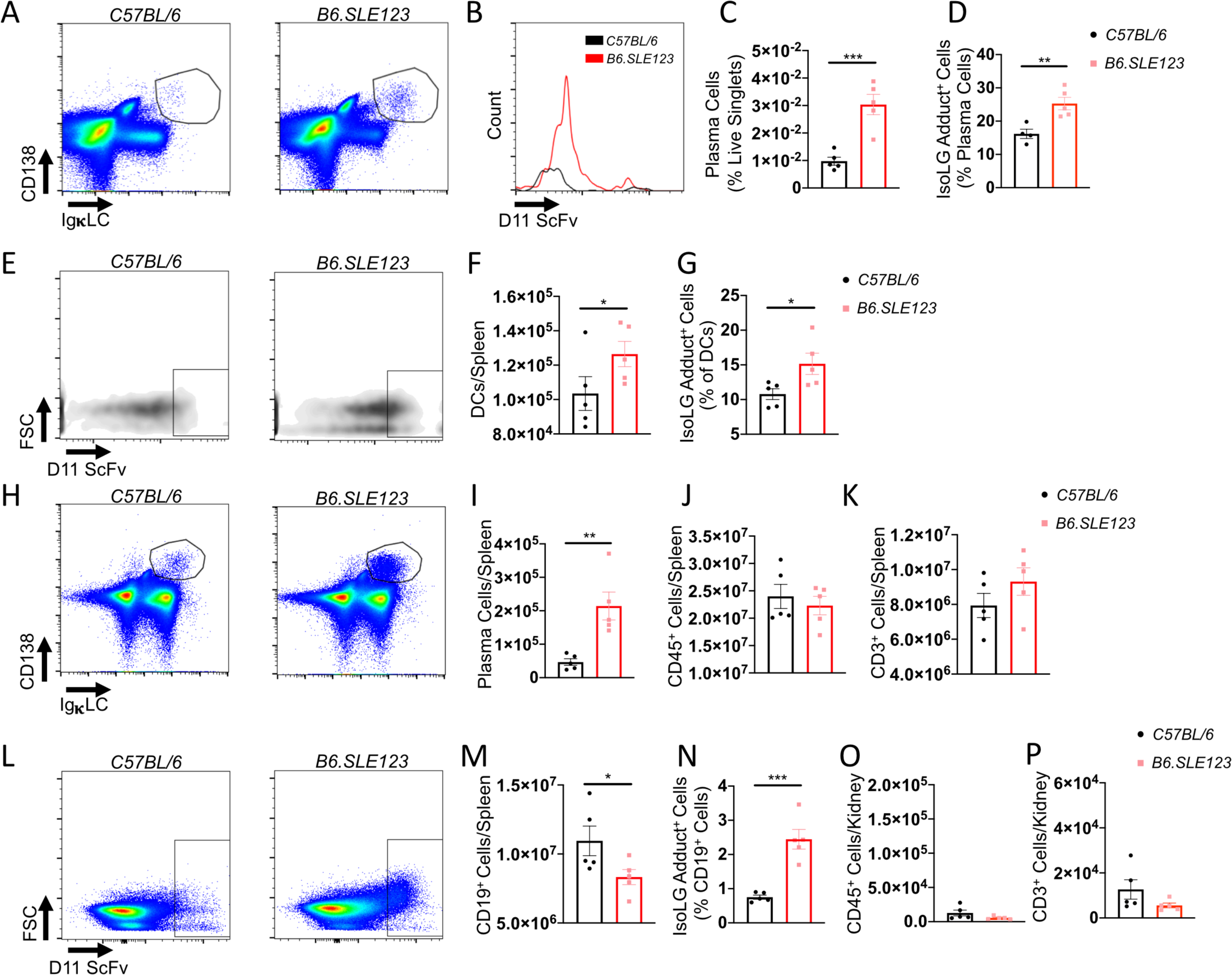
IsoLG adduct accumulation and immune dysfunction in 7-week old *B6.SLE123* Mice. Single cell suspensions were prepared from freshly isolated mouse tissue via enzymatic digestion and mechanical dissociation. Live cell singlets were analyzed. Representative FACS plots are presented for **(A)** bone marrow plasma cells. **(B)** A histogram of isoLG adduct accumulation in bone marrow plasma cells measured using the single chain antibody D11 ScFv. **(C)** Quantitation of bone marrow plasma cells. **(D)** isoLG adduct containing plasma cells. **(E)** Representative density plots of splenic dendritic cell isoLG adduct accumulation. **(F)** Quantitation of dendritic cells in spleen and **(G)** isoLG adduct containing DCs in spleen. **(H)** Representative FACS plots of plasma cells in spleen. Quantitation of **(I)** splenic plasma cells **(J)** splenic CD45^+^ cells and **(K)** splenic CD3^+^ T cells. **(L)** Representative FACS plots of isoLG adduct accumulation in CD19^+^ B cells in spleen. Quantitation of **(M)** splenic CD19^+^ B cells **(N)** isoLG adduct containing CD19^+^ B cells in spleen **(O)** kidney CD45^+^ cells and **(P)** kidney CD3^+^ cells. Data were analyzed using Student’s T-test (*n* = 5, *\*P* < 0.05, *\*\*P* < 0.01, *\*\*\* P* < 0.001, *\*\*\*\* P* < 0.0001).

**Figure 2:**
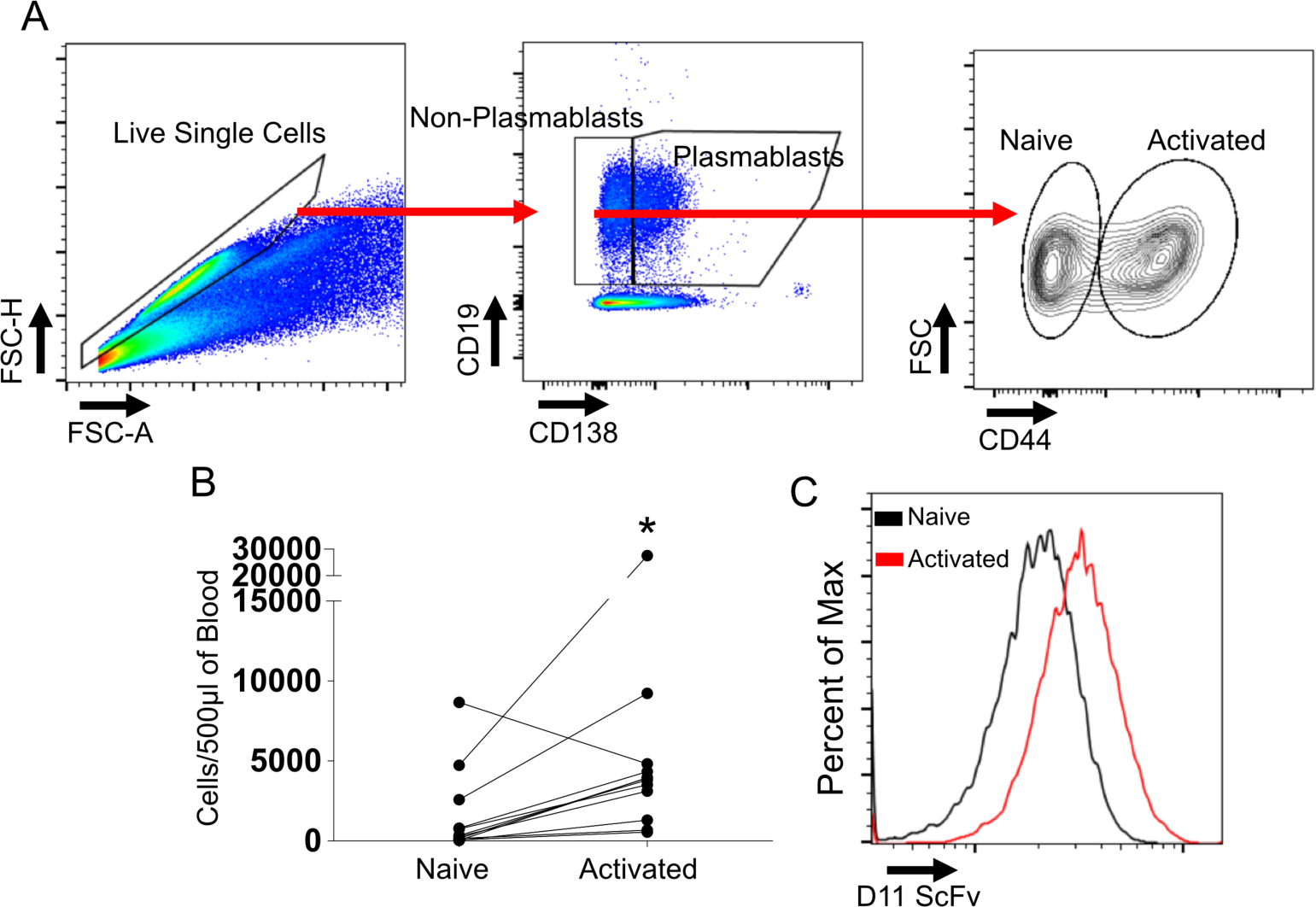
IsoLG adducts are enriched in peripheral activated B cells of SLE prone mice. Cells were isolated at the time of sacrifice from 32-week old *B6.SLE123* mice. Activated B cells in peripheral blood exhibit augmented isoLG adduct accumulation compared to naive B cells. **(A)** Gating strategy to define activated vs naive B cells. Gating was performed on non-plasmablast B cells. **(B)** IsoLG adduct containing cells in peripheral blood of *B6.SLE123* and *C57BL.6* mice in peripheral blood is quantitated. Data were analyzed with a paired T-test (*n* = 12, ^*\**^*P* < 0.05) **(C)** Representative histograms of isoLG adduct levels in activated vs naive B cells.

### Scavenging of isoLGs reduces isoLG adduct accumulation in murine SLE

To further investigate a role of isoLG adducts in SLE, *B6.SLE123* animals were treated with 2-HOBA beginning at 7 weeks of age. Animals were sacrificed at 32 weeks of age and peripheral blood, spleen, and bone marrow cells were harvested from *B6.SLE123* and control *C57BL/6* mice and flow cytometry was performed. Ly6c is a marker of monocyte activation and denotes a pro-inflammatory phenotype (24). Interestingly, intracellular isoLG adducts within splenic DCs (Figure 3 A-C), peripheral blood total and Ly6c^+^ monocytes (Figure 3 D-F), and bone marrow plasma cells (Figure 3 G and H) were markedly increased in *B6.SLE123* mice compared to control *C57BL/6* mice whereas treatment with 2-HOBA attenuated this accumulation in *B6.SLE123* mice.

**Figure 3:**
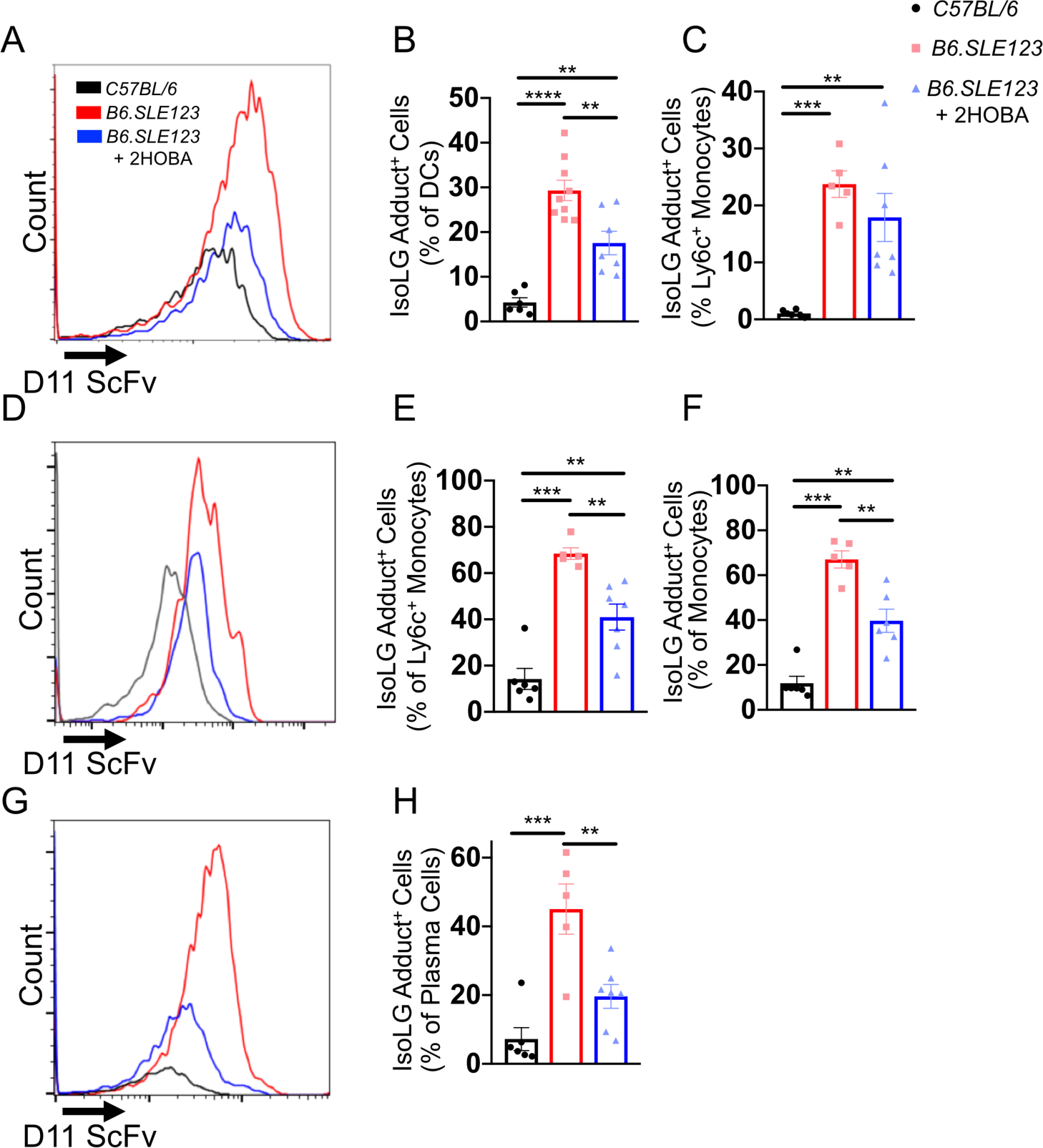
IsoLG adducts are enriched in SLE prone mice and are efficiently scavenged by 2-HOBA. Cells were isolated at the time of sacrifice from 32-week old *B6.SLE123* mice. Single cell suspensions were prepared from freshly isolated mouse tissue via enzymatic digestion and mechanical dissociation. Live cell singlets were analyzed. **(A)** Representative histograms revealing isoLG adduct enrichment and efficient scavenging by 2-HOBA in DCs from spleens. Quantitation of isoLG adduct containing cells in **(B)** splenic DCs **(C)** splenic Ly6c^+^ monocytes **(D)** Representative histogram from peripheral blood Ly6c^+^ monocytes and quantitation of **(E)** isoLG adduct containing peripheral blood Ly6c^+^ monocytes **(F)** peripheral blood total monocytes. **(G)** representative histogram and **(H)** quantitation of isoLG adduct containing bone marrow plasma cells. Data were analyzed using 1-way ANOVA (*n* = 5-9, *\*P* < 0.05, *\*\*P* < 0.01, *\*\*\* P* < 0.001, *\*\*\*\* P* < 0.0001).

### Scavenging of isoLG adducts reduces splenic myeloid and lymphoid expansion in SLE

SLE is characterized by robust expansion of multiple cell types including myeloid cells, T cells, and plasma cells. *B6.SLE123* mice exhibit a significant increase in spleen size and an expansion of myeloid and lymphoid cells. Spleens from 2-HOBA treated *B6.SLE123* were reduced in size (Figure 4A). Reduction in spleen size was also evidenced by a reduction in total cell number as measured by flow cytometry (Figure 4B). This was accompanied by a reduction in CD45^+^ cells (Figure 4C). Specifically, DCs, CD3^+^, CD4^+^, CD8^+^, and CD19^+^, cells were significantly reduced in spleens of *B6.SLE123* mice treated with 2-HOBA (Figure 4 D-J). These data suggest that scavenging of isoLGs reduces the expansion of multiple cell types in SLE.

**Figure 4:**
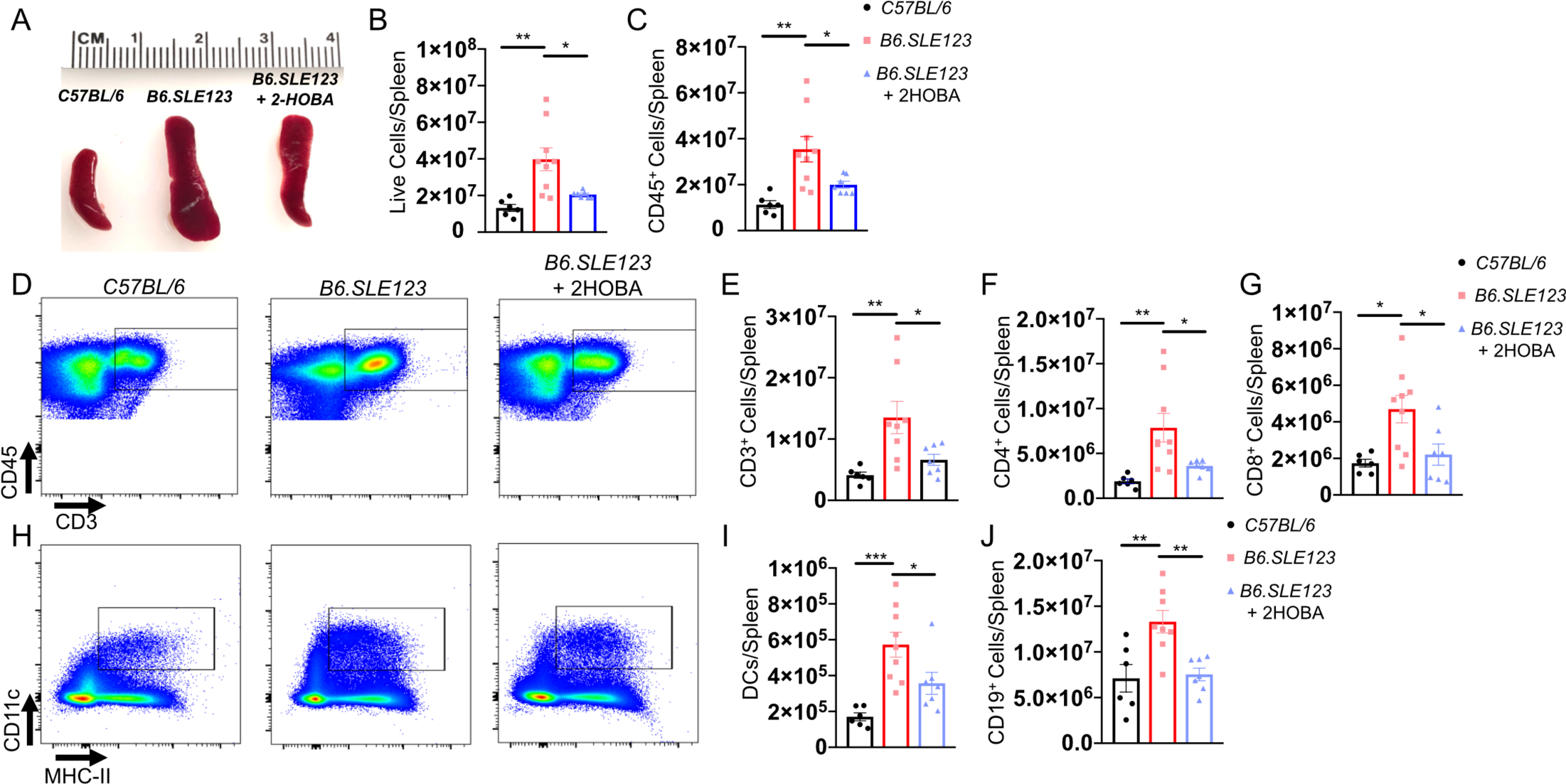
Scavenging of isoLG reduces splenic myeloid and lymphoid expansion in a mouse model of SLE. Cells were isolated at the time of sacrifice from 32-week old *B6.SLE123* mice. Single cell suspensions were prepared from freshly isolated mouse tissue via enzymatic digestion and mechanical dissociation. Live cell singlets were analyzed. **(A)** Representative spleens revealing a reduction in spleen size from *B6.SLE123* mice treated with 2-HOBA. Quantitation of **(B)** Live cells and **(C)** CD45^+^ cells are presented. **(D)** Representative FACS plots for CD3^+^ T cells. Quantitation of **(E)** CD3^+^ T cells **(F)** CD4^+^ T cells **(G)** CD8^+^ T cells. Representative FACS plots for **(H)** CD11c^+^/MHCII^+^ DCs. Quantitation of **(I)** DCs **(J)** and CD19^+^ B cells. Data were analyzed using 1-way ANOVA (*n* = 6-9, *\*P* < 0.05, *\*\*P* < 0.01, \*\*\**P* < 0.001).

### SLE-associated hypertension is attenuated by 2-HOBA

In the DOCA-salt and angiotensin II mouse models of hypertension, the isoLG scavenger 2-HOBA significantly attenuates blood pressure elevation and end-organ damage (20). This agent does not react with superoxide, peroxynitrite or hydrogen peroxide. To determine the importance of isoLG adduct accumulation in SLE associated hypertension, we implanted telemeters at 30-weeks of age in a subset of *B6.SLE1213* mice, treated or not with 2-HOBA. As evidenced by telemetry recordings, systolic, diastolic, and mean arterial blood pressures were significantly reduced in *B6.SLE123* animals treated with 2-HOBA when compared to *B6.SLE123* animals treated with vehicle (Figure 5 A-C). These data indicated that scavenging of isoLG improves arterial hypertension in SLE.

**Figure 5:**
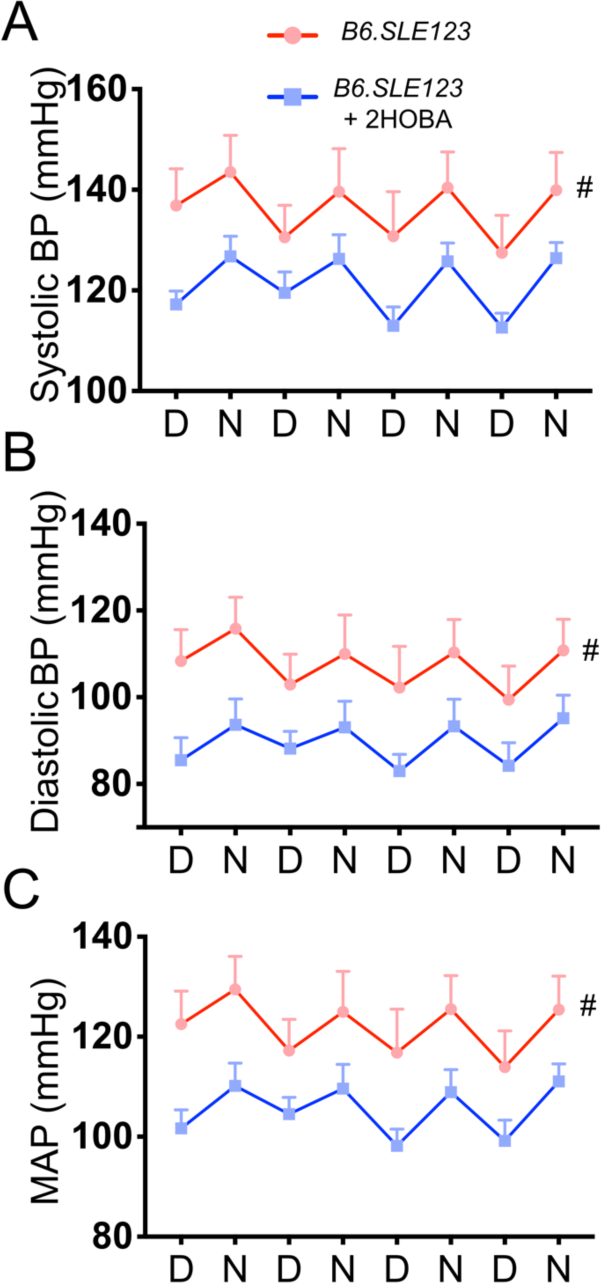
IsoLG scavenging attenuates hypertension in a mouse model of SLE. Radiotelemeters were implanted in 30-week old animals. Measurements were made prior to sacrifice at 32-weeks old. Average measurements over a three-day period. Day (D) and night (N) cycles are represented for **(A)** systolic, **(B)** diastolic, **(C)** mean arterial pressures. Blood pressure was analyzed using 2-way ANOVA (*n*=6-7, ^*#*^ *P* < 0.001).

### Improvement in SLE associated renal injury and function with 2-HOBA

Patients with hypertension and SLE often exhibit abnormal sodium excretion and augmented albuminuria; moreover, glomerular injury secondary to immune complex deposition occurs in many patients with SLE (25, 26). Using flow cytometry of single cell preparations of kidney homogenates, we observed a marked reduction in inflammation as evidenced by a reduction in renal CD45^+^ cells, CD3^+^ T cells, and CD4^+^ T cells in *B6.SLE123* animals treated with 2-HOBA (Figure 6 A-F). We did not observe a significant reduction in CD8^+^ T cells (Figure 6 G). To examine the effects of isoLG scavenging on renal function and injury, *B6.SLE123* animals treated with and without 2-HOBA were injected with 4% normal saline at a volume equal to 10% of their body weight as previously described (6, 7, 27). Urine volume was measured over the subsequent 4 hours. The volume of urine excreted compared to total body weight was significantly reduced in *B6.SLE123* animals compared to healthy controls. Treatment with 2-HOBA improved urine excretion (Figure 7A). The albumin to creatinine ratio was significantly elevated in *B6.SLE123* animal as was urinary excretion of NGAL, a marker of glomerular injury. These parameters were also attenuated with 2-HOBA treatment (Figure 7B-C). Jones silver stain of paraffin-embedded kidneys revealed that 2-HOBA treatment also reduced glomerular hypercellularity, wire loop lesions, and immune complex deposition (Figure 7D, E-G). These findings suggest that isoLGs contribute to renal inflammation, injury, and immune complex deposition in SLE.

**Figure 6:**
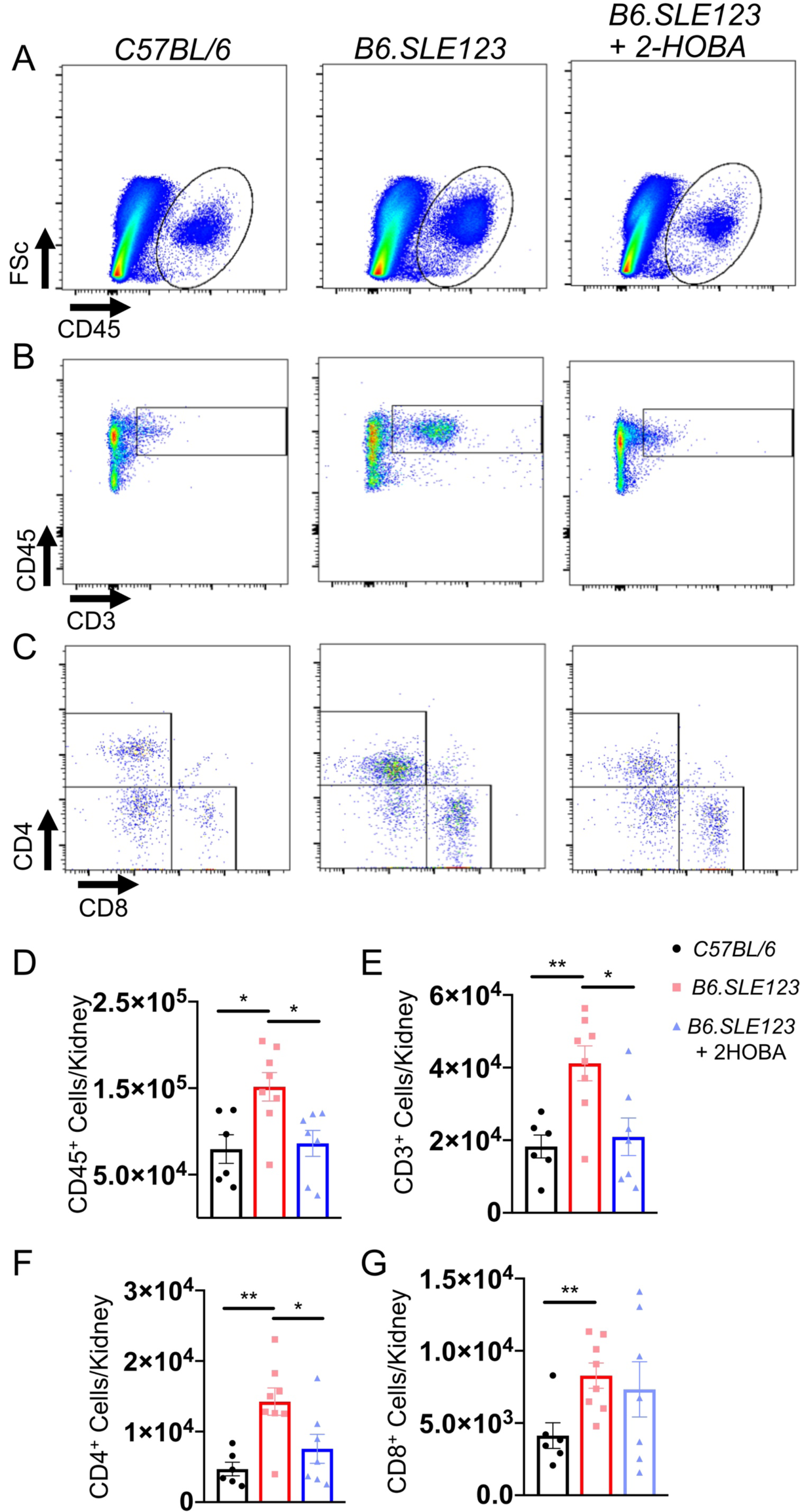
Scavenging of isoLG attenuates renal inflammation in a mouse model of SLE. . Cells were isolated at the time of sacrifice from 32-week old *B6.SLE123* mice. Single cell suspensions were prepared from freshly isolated mouse tissue via enzymatic digestion and mechanical dissociation. Live cell singlets were analyzed. Representative FACS plots are presented for **(A)** CD45^+^ total leukocytes **(B)** CD3^+^ T cells **(C)** CD4^+^and CD8^+^ T cells. Quantitation of **(D)** CD45^+^ leukocytes **(E)** CD3^+^ T cells **(F)** CD4^+^ and **(G)** CD8^+^ T cells are represented. Data were analyzed using 1-way ANOVA (*n* = 6-8, *\*P* < 0.05, *\*\*P* < 0.01, \*\*\**P* < 0.001, \*\*\*\**P* < 0.0001).

**Figure 7:**
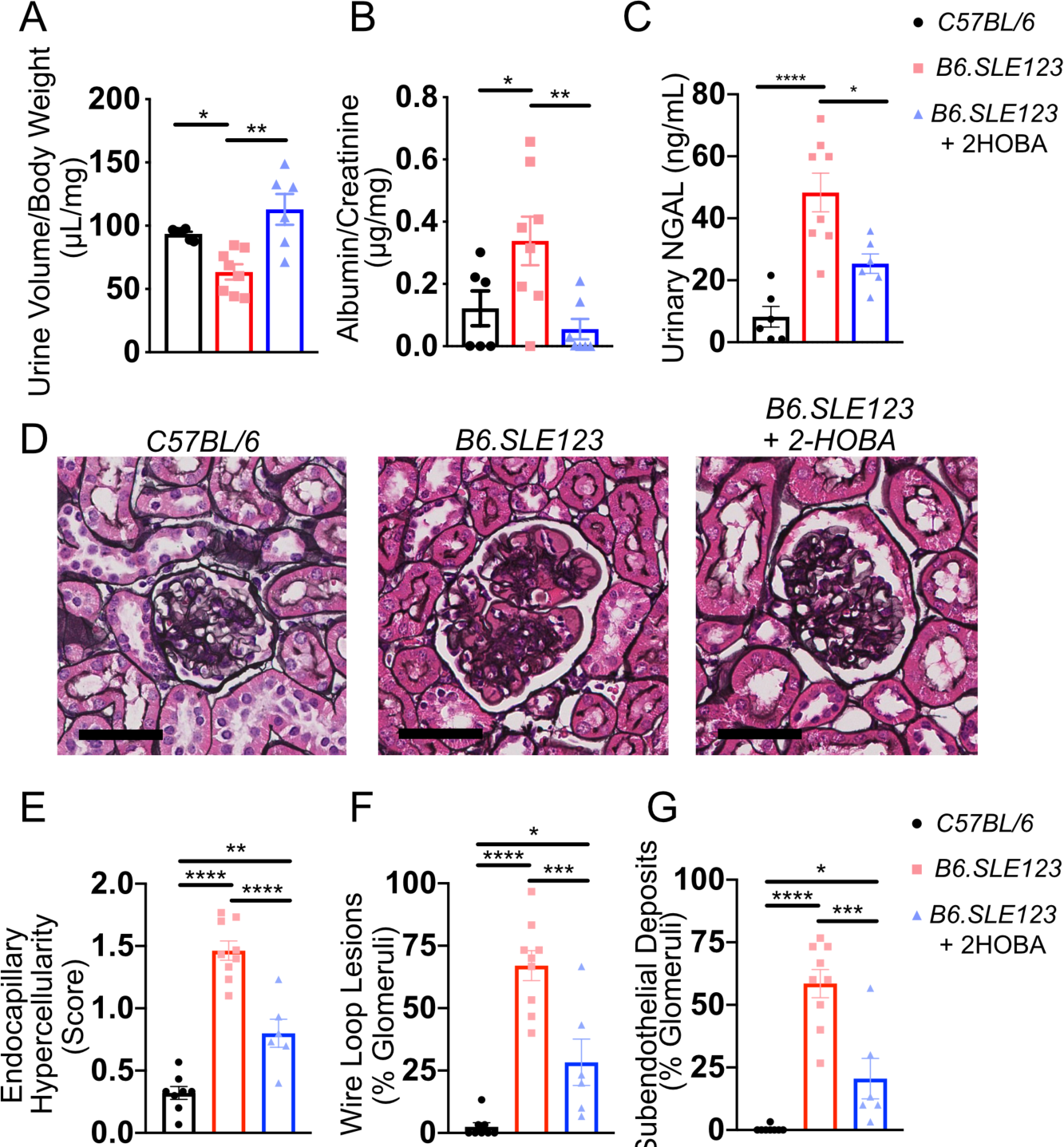
Renal dysfunction in a mouse model of SLE is attenuated by 2-HOBA. Urine studies were performed on 31-week old animals. Animals were sacrificed at 32-week old and kidneys were stained with Jones’ **(A)** Mice received intraperitoneal injection of 4% normal saline at 10% of body weight and urine output was measured after 4-hours. **(B)** Spot urine albumin/creatine ratio. **(C)** Spot urinary NGAL which represents glomerular injury was quantitated. Kidneys were sectioned and stained with Jones’ silver stain and **(D)** representative glomeruli are presented (bar = 40μm). Kidneys were scored for **(E)** severity of endocapillary hypercellularity **(F)** presence of wire loop lesions and **(G)** subendothelial deposits. Data were analyzed using 1-way ANOVA (*n* = 6-8, *\*P* < 0.05, *\*\*P* < 0.01, \*\*\**P* < 0.001, \*\*\*\**P* < 0.0001).

### Scavenging of isoLG reduces bone marrow plasma cell expansion and anti-dsDNA antibody titers

SLE is associated with an expansion of plasma cells and an elaboration of autoantibodies. Autoreactive plasma cells are believed to play an important role in SLE by generating autoantibodies that lead to the creation of immune complexes which deposit in peripheral tissues (28). Flow cytometry revealed a significant accumulation of spleen and bone marrow plasma cells in *B6.SLE123* animals, which is attenuated by treatment with 2-HOBA (Figure 8A-C). Plasma cells are responsible for secretion of autoantibodies in SLE. We found that *B6.SLE123* animals treated with 2-HOBA exhibit markedly lower plasma anti-dsDNA antibody and total IgG antibody titers (Figure 8D-E). These data demonstrate that scavenging of isoLGs with 2-HOBA attenuates plasma cell accumulation and autoantibody production.

**Figure 8:**
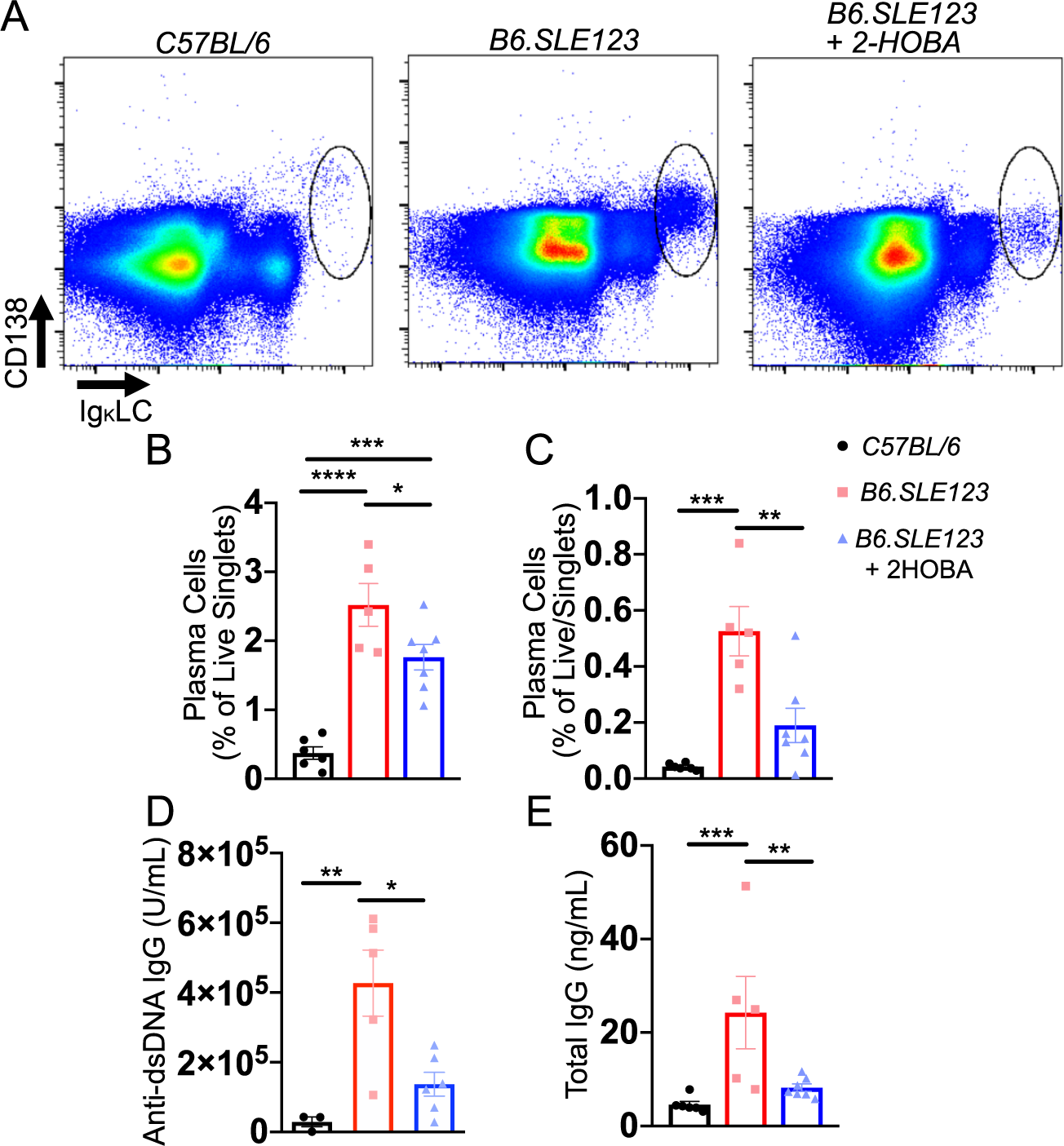
Scavenging of isoLG reduces plasma cell expansion in spleen and bone marrow and reduces total IgG and anti-dsDNA antibody titers in a mouse model of SLE. Cells were isolated at the time of sacrifice from 32-week old *B6.SLE123* mice. Single cell suspensions were prepared from freshly isolated mouse tissue via enzymatic digestion and mechanical dissociation. Live cell singlets were analyzed. Live singlets were analyzed. **(A)** Representative FACS plots displaying CD138^+^ and intracellular Igκ-light-chain^+^ plasma cells from bone marrow. Quantitation of plasma cells as a percentage of CD45^+^ cells are represented for **(B)** spleen and **(C)** bone marrow. **(D)** Anti-dsDNA IgG antibody and **(E)** total IgG at 1:100K dilution was quantified from plasma using ELISA. Data were analyzed by 1-way ANOVA (*n* = 5-7, ^*\**^*P* < 0.05, ^*\*\**^*P* < 0.01, ^*\*\*\**^*P*<0.001, ^*\*\*\*\**^*P*<0.0001).

### 2-HOBA reduces blood pressure, renal inflammation, and plasma cell accumulation in the *NZBWF1* mouse model of SLE

To determine if isoLG adducts also contribute to disease in this independent model of hypertension, *NZBWF1* mice were treated with continuous 2-HOBA beginning at 7-weeks of age. Importantly, untreated animals displayed a marked accumulation of isoLG within splenic dendritic cells and bone marrow plasma cells, similar to that observed in the *B6.SLE123* model, and 2-HOBA efficiently prevents isoLG adduct formation (Figure 9 A-D). Systolic, diastolic, and mean arterial pressure was markedly reduced in the *NZBWF1* animals treated with 2-HOBA (Figure 9 E-G) as measured by radiotelemetry. Moreover, renal immune cell and T cell infiltration was markedly attenuated by treatment, similar to the *B6.SLE123* model (Figure 9 H,I). Total splenic T cells and DCs were likewise reduced (Figure 9 J,K). Bone marrow plasma cell accumulation was reduced with 2-HOBA treatment (Figure 9 L). Similarly, as observed in the *B6.SLE123* model. urinary NGAL excretion was attenuated by treatment with 2-HOBA in *NZBWF1* mice (Figure 9 M). These data confirm that our findings in the *B6.SLE123* model in an independent model of SLE.

**Figure 9:**
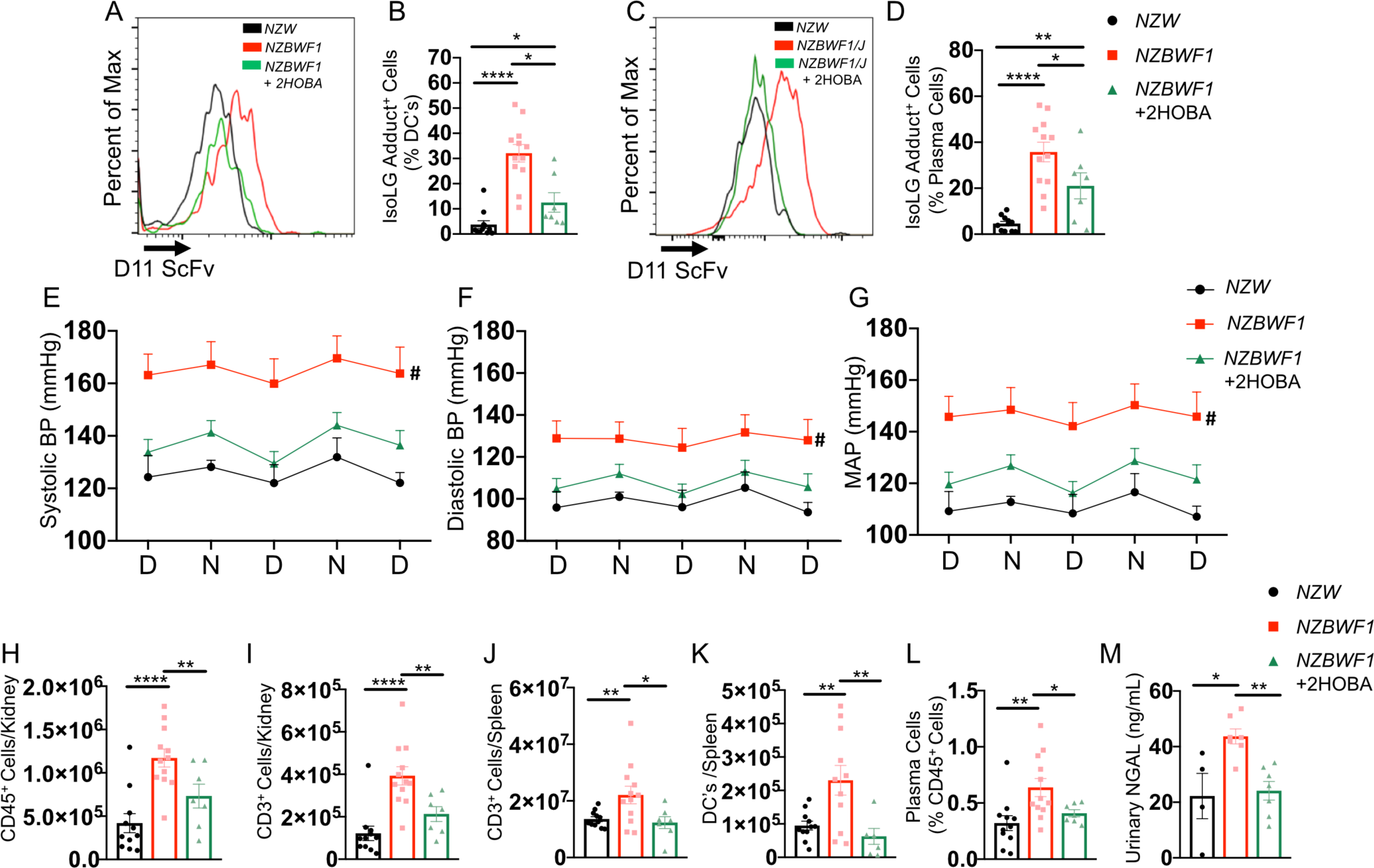
IsoLG Scavenger 2-HOBA reduces blood pressure systemic autoimmunity in the *NZBWF1* mouse model of SLE. Animals were sacrificed at 32-weeks old and single cell suspensions were prepared from spleen, kidney, and bone marrow and analyzed by flow cytometry. **(A)** Representative histogram displaying the distribution of isoLG adduct containing splenic DCs. **(B)** Quantitation of isoLG adduct containing splenic DCs. Representative histogram displaying the distribution of isoLG adduct containing bone marrow plasma cells. Quantitation of isoLG adduct containing bone marrow plasma cells. Radiotelemeters were implanted in 30-week old mice and Blood pressure was measured over a three day period prior to sacrifice at 32-weeks old. Day and night cycles are represented for **(E)** systolic **(F)** diastolic **(G)** mean arterial pressure. Flow cytometry was performed for quantitation of **(H)** kidney CD45^+^ and **(I)** kidney CD3^+^ T cells **(J)** splenic CD3^+^ T cells **(K)** splenic DCs and **(L)** bone marrow plasma cells are represented. **(M)** Urinary NGAL was quantitated by ELISA. Data from B,D, H-M were analyzed by 1-way ANOVA (*n* = 7-12, ^*\**^*P* < 0.05, ^*\*\**^*P* < 0.01, ^*\*\*\**^*P*<0.001, ^*\*\*\*\**^*P*<0.0001). Blood pressure was analyzed using 2-way ANOVA (*n*=5-9, ^*#*^ *P* < 0.001).

### Serum from the *B6.SLE123* and *NZBWF1/J* murine models reacts with IsoLG adducted protein

Previous studies have described the autoantigenicity of oxynonenol- and hydroxynonenol-adducted peptides in SLE (29). We hypothesized that isoLG-adducted protein would react with serum from mice with SLE. To test this hypothesis, we analyzed serum reactivity to isoLG adducts by ELISA and immunoblot. We developed a capture ELISA to detect the presence of isoLG adduct-specific IgG antibodies in serum from mice (Figure 10A). Interestingly, both the *B6.SLE123* and *NZBWF1* models exhibited marked reactivity of serum with isoLG adducts with the *NZBWF1* animal model displayed greater variability. Importantly, scavenging of isoLG with 2-HOBA prevented accumulation of these antibodies (Figure 10B-C).

**Figure 10:**
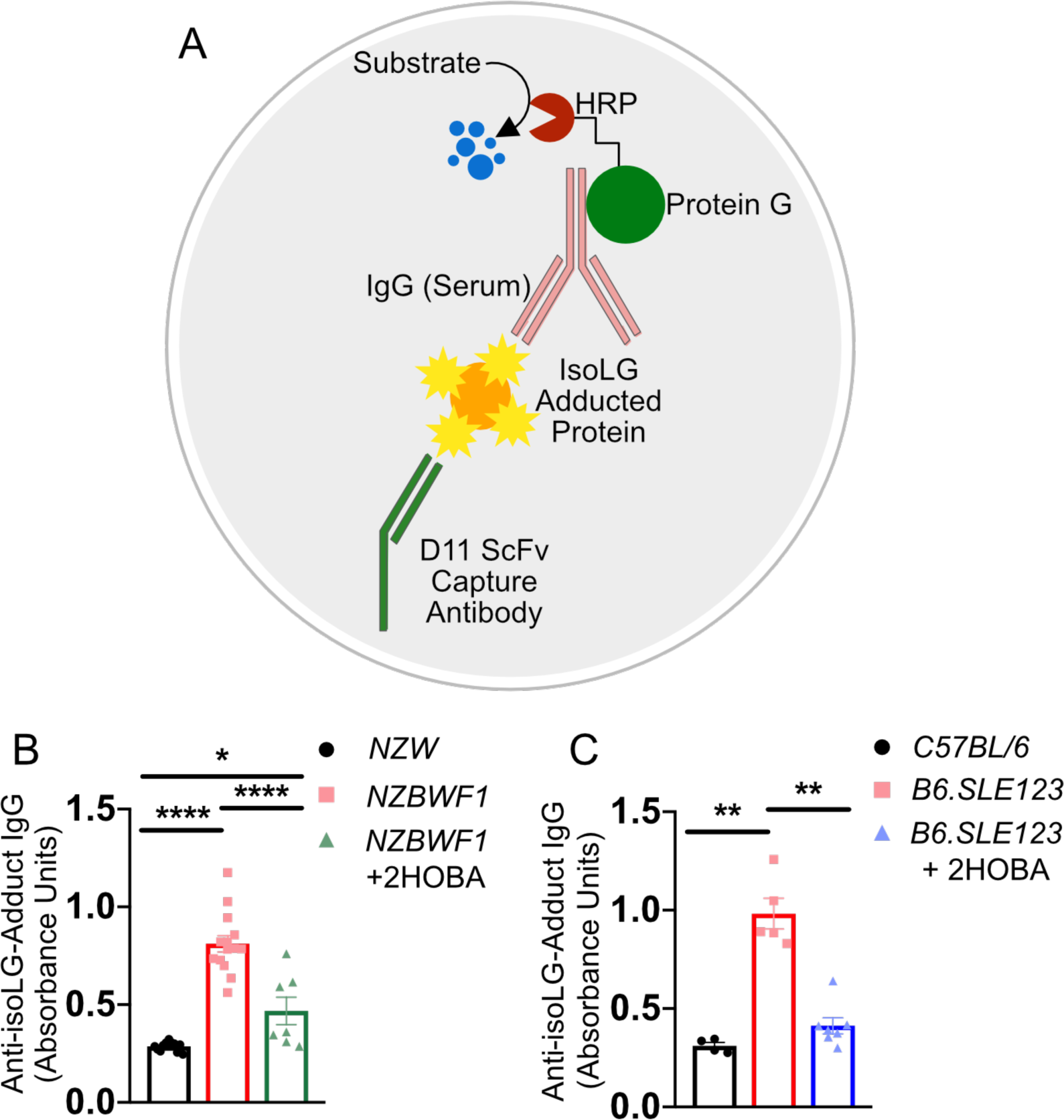
IsoLG adducts react with serum from SLE prone mice. Serum was collected from mice at the time of sacrifice at 32-weeks old. Antibodies against isoLG adducts were determined by ELISA from serum. **(A)** Model of capture assay to detect anti-isoLG adduct IgG. The D11 ScFv single chain antibody is bound to a plate. Kidney protein was adducted with isoLG *in vitro* and incubated with D11 ScFv. This is followed by serum from lupus prone mice. The presence of IgG was detected utilizing a protein-G horse radish peroxidate (HRP) conjugate. Anti-isoLG adduct IgG was detected in **(B)** *NZBWF1* mice treated with vehicle or 2-HOBA compared to *NZW* controls and **(C)** *B6.SLE123* mice treated with vehicle or 2-HOBA compared to *C57BL/6* controls Data were analyzed with 1-way ANOVA (*n* = 4-7 for *B6.SLE123, n =* 7-14 for *NZBWF1*).

### IsoLG adducts are enriched in monocytes of patients with SLE

To determine if isoLG adducts accumulate in myeloid cells of humans with SLE, we employed flow cytometry to examine peripheral blood mononuclear cells (PBMC) from 11 subjects with SLE and 10 age-sex matched healthy controls as previously described (20, 30). SLE subjects were recruited from rheumatology clinic. Age and sex matched healthy control subjects were recruited by local advertising. The majority of SLE patients were female and all were positive for antinuclear antibodies. 64% of SLE patients were on ACE inhibitor therapy (Table 1). Additional patient characteristics are shown in Table 1. IsoLGs were identified in cells expressing CD11c, CD14, CD86, and CD83. IsoLG adducts were enriched in CD11c^+^ and CD14^+^ monocytes in addition to CD11c^+^ monocytes that are positive for the activation marker CD86 (Figure 11 A-F). CD45^+^ Cells positive for the activation marker CD83 exhibited no significant difference in isoLG adduct accumulation (Figure 11 G). In addition to flow cytometry, we sorted monocytes from an additional set of 6 SLE patients and 4 matched control subjects and quantitated isoLG-lysine adducts by mass spectrometry. This confirmed an 8-fold enrichment of isoLG adducts in monocytes of SLE subjects compared to controls (Figure 11 H-I, Supplemental Table 1). Thus, the accumulation of isoLG protein adducts is markedly increased in circulating myeloid cells of humans with SLE. These levels are similar to those observed in humans with essential hypertension and suggest a potential common mechanism in the initiation of essential hypertension and SLE (20). In experimental hypertension, the formation of isoLG adducts within APCs is accompanied by augmented superoxide production (31). To determine if superoxide production is augmented in monocytes from SLE patients, we measured the formation of 2-hydroxyethidium from dihydroethidium in isolated monocytes from a third cohort of 6 SLE subjects and 4 matched controls and observed an approximately nine-fold increase in 2-hydroxyethidium in SLE monocytes compared to controls (Figure 11 J-K, Supplemental Table 2).

**Table 1:** Clinical features of SLE patients and controls studied for with flow cytometry. Data on musculoskeletal involvement and anti-dsDNA antibody data unavailable in 1 subject. SLICC data unavailable in 2 subjects

|  | SLE Median [IQR]<br>or # (%) | Control Median [IQR]<br>or # (%) |
| --- | --- | --- |
| **Demographics** |  |  |
| Age, years | 39 [27, 54] | 34.5 [31, 64] |
| Race, # Caucasian | 6 (55) | 6 (60) |
| Ethnicity, # Hispanic | 3 (27) | 0 (0) |
| Sex, # female | 10 (91) | 7 (70) |
| Blood Pressure (Systolic) | 123.5 (108-150) | 129 (118-137) |
| Blood Pressure (Diastolic) | 75 (51-93) | 79 (63-89) |
| BMI | 24.5 (17-40.2) | 25.4 (22.2-36.7) |
| Creatinine | 0.84 (0.76-0.87) | N/A |
| **SLE Manifestation** |  |  |
| Mucocutaneous, # ever | 9 (82) | N/A |
| Musculoskeletal, # ever | 7 (64) | N/A |
| Serositis, # ever | 8 (72) | N/A |
| Hematologic, # ever | 6 (55) | N/A |
| Renal, # ever | 6 (55) | N/A |
| Anti-dsDNA positivity, # | 6 (60) | N/A |
| Low complement, # ever | 6 (55) | N/A |
| SLEDAI, score | 2 [2, 4] | N/A |
| SLICC, score | 1 [0, 1] | N/A |
| Hypertension | 2 (18) |  |
| **Current medication use** |  |  |
| Hydroxychloroquine, # | 10 (91) | N/A |
| Mycophenolate, # | 6 (55) | N/A |
| Corticosteroids, # | 5 (45) | N/A |
| ACE Inhibitors | 7 (64) |  |

**Figure 11:**
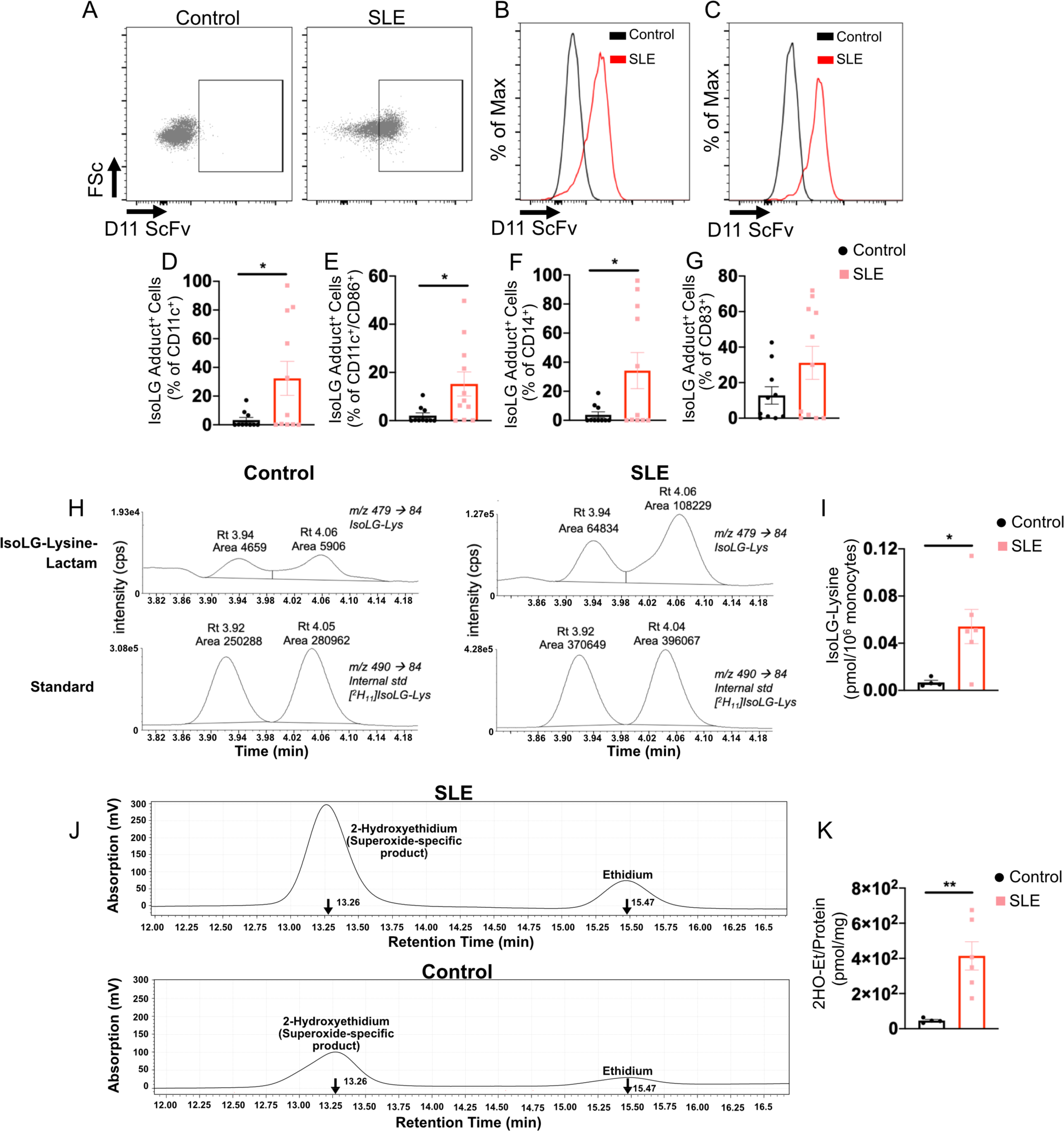
IsoLG adducts are enriched in monocytes of patients with SLE. **(A)** Representative FACS plots displaying isoLG adduct containing CD11c^+^ PBMCs from a representative control and SLE subject. Representative histograms displaying the distribution of isoLG adducts in **(B)** CD11c^+^ and **(C)** CD11c^+^/CD86^+^ cells. Quantitation of IsoLG adduct containing cells as a percentage of **(D)** CD11c^+^, **(E)** CD11c^+^/CD86^+^, **(F)** CD14^+^ and **(G)** CD83^+^ cells. For D-G data were analyzed using Student’s T-test (*n* = 10-11, *\*P* < 0.05). **(H)** Stable isotope dilution multiple reaction monitoring for mass spectrometry analysis of isoLG-lysine-lactam adduct in DCs. Representative LC/MS chromatographs from a representative patient is shown. The top row shows multiple reaction monitoring chromatographs for isoLG lysine lactam (isoLG-Lys) in sample, while the bottom row shows multiple reaction monitoring chromatograph for [13C615N2] internal standard for the same samples. **(I)** Quantitation of isoLG-lysine in monocytes from a subset of SLE subjects and controls. **(J)** Monocytes from a SLE patients and controls were sorted. Superoxide was detected using HPLC to monitor conversion of dihydroethidium to the superoxide oxidation adduct 2-hydroxyethidium (2-HO-Et) and ethidium. **(K)** Quantitation of 2-HO-Et from SLE subjects and controls. For I and K, comparisons were made with a Student’s T-test (*n* = 4-6, *\*P* < 0.05).

## DISCUSSION

In this study, we define a previously unknown role of isoLGs and their protein adducts in the pathogenesis of SLE and its associated hypertension and renal disease. We identified an accumulation of isoLG adducts in monocytes of patients with SLE and in antigen presenting cells of mice with SLE and defined novel potential functional roles of isoLG adduction. The present study suggests that prior to the onset of overt disease, isoLG adduct accumulation occurs in DCs, B cells, and plasma cells of *B6.SLE123* animals. Moreover, continuous scavenging of isoLG initiated early in the disease course attenuates disease activity. These effects are pleiotropic and include a reduction in blood pressure, myeloid and lymphoid expansion, and autoantibody production. Given the initial changes observed in young SLE prone mice in addition to the wide-ranging effects of 2-HOBA, it is plausible that isoLG adduction is an initiating step in SLE (Figure 12).

**Figure 12:**
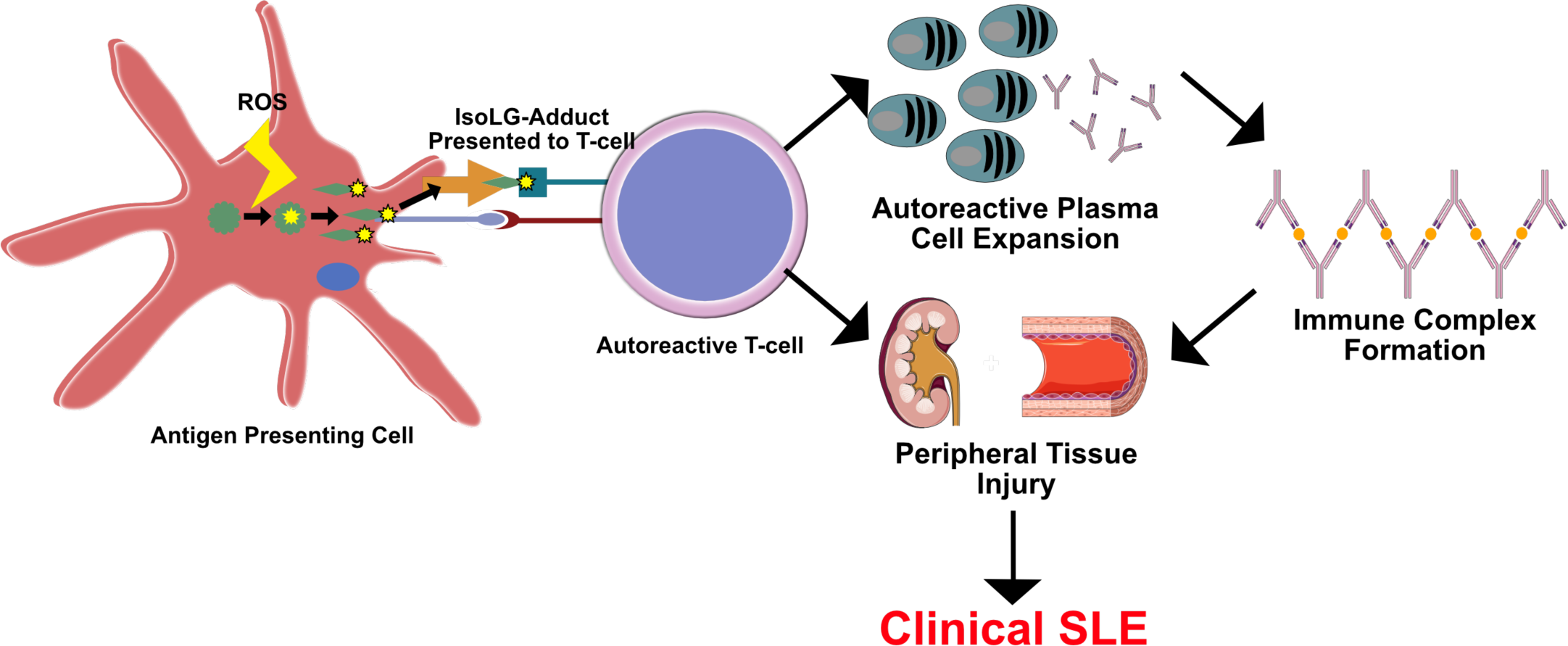
Paradigm of isoLG-induced activation of autoreactive T cells in the development of SLE. Within an antigen presenting cell, intracellular ROS results in formation of reactive isoLG (yellow stars) which adduct to protein (Green stars). Intracellular proteins are processed and presented on the cell surface to autoreactive T cells which then migrate to peripheral tissues and drive development of autoreactive plasma cells. Plasma cells secerete antibodies that form immune complexes and contribute to renal and peripheral tissue injury and dysfunction leading to clinical SLE.

For SLE patients, hypertension is an important and powerful risk factor for the development of numerous comorbidities. Importantly, women with SLE exhibit an increased prevalence of hypertension when compared to controls (32). Up to 30% of patients with proliferative nephritis associated with SLE develop end-stage renal disease (ESRD) (33). Importantly, the presence of hypertension at initial diagnosis of nephritis is associated with rapid development of ESRD (34). Hypertension is often present in the absence of nephritis in SLE patients, suggesting the importance of an early intervention strategy prior to the development of renal disease (35). In addition to renal disease, SLE patients exhibit a markedly increased risk of cardiovascular disease (32, 36). Specifically, patients with SLE exhibit accelerated development of atherosclerosis and coronary heart disease (CHD) (37-40).

In addition to traditional risk factors, inflammation and associated clinical factors contribute to the development of CHD and ESRD in SLE patients (32). Oxidative injury has been implicated in SLE pathogenesis and the development of SLE-associated comorbidities (41). In the *NZBWF1* model of SLE, renal cortical oxidative stress is increased and treatment with etanercept, an anti-TNF*α* therapy, attenuated both hypertension and oxidative stress (42). As mentioned above, a 4-week treatment of *NZBWF1* animals beginning at 30 weeks of age with the antioxidant tempol and the NADPH oxidase inhibitor apocynin attenuated hypertension (17). This therapy did not, however, alter disease progression as measured by anti-dsDNA antibody titers. These data are supported by numerous studies that suggest a role of ROS in the pathogenesis of SLE (12, 25, 41). The current study confirms an increase in superoxide production within monocytes of patients with SLE and supports a role of downstream isoLGs as a mechanism for systemic immune activation and hypertension in SLE.

Accumulation of isoLG adducts within CD11c^+^ and CD14^+^ monocytes is observed with concomitant accumulation within the activated CD11c^+^/CD86^+^ subpopulation. Importantly, we found a wide variation in the presence of IsoLG adduct accumulation among patients with SLE and also found that 3 of 4 SLE patients examined exhibit serum reactivity with isoLG adducted protein. Patients with SLE exhibit significant disease heterogeneity (26). The age of onset, clinical manifestations, and serologic abnormalities are vary markedly (43, 44). It is conceivable that isoLG adduct accumulation occurs in a subset of SLE patients, leading to specific disease manifestations and organ involvement. The presence of isoLG adducts might serve as a biomarker to indicate patients that would benefit from isoLG scavenging therapy. Future studies should focus on identification of this subpopulation of patients.

In prior studies we have shown that exposure of DCs to tert-butyl-hydroperoxide, a lipid soluble peroxide molecule, leads to formation of intracellular isoLG adducts. Adoptive transfer of these oxidatively modified cells primes hypertension to a generally subpressor dose of ang II in mice (20). These data suggest that isoLG formation in APCs such as DCs and monocytes play an important role in hypertension. A similar mechanism is likely operative in SLE whereby isoLG adducts displayed by professional APCs result in activation of an inflammatory cascade resulting in autoreactive CD8^+^ and CD4^+^ T cell activation (Figure 12). Prevention of this initial step may attenuate this process and thus systemic autoimmunity and hypertension.

Importantly, the reactivity of isoLG adducted protein with serum from SLE prone mice suggests that isoLG adducts are recognized as autoantigens that lead to the production of autoantibodies. In SLE, the induction of autoantibody production is affected by dysfunctional Tfh cells (45-47). However, dysregulation of B cells in SLE is also cell-autonomous. In healthy individuals, autoreactive B cells are limited due to multiple mechanisms of tolerance induction, including receptor editing, apoptosis, and anergy (48). These B cells do not reach the germinal center where activation occurs (48). In patients with SLE, autoreactive B cells mature from non-autoreactive precursors and memory B cells (49). In addition to activation, B cell receptor recognition of soluble or tethered antigens induces endosomal and lysosomal compartment formation and internalized antigens are processed to form MHC class II-peptide complexes (50). It is conceivable that, in addition to activation by isoLG adduct presenting APCs and isoLG adduct specific CD4^+^ T cells, autoreactive B cells accumulate isoLG adducts during endocytosis. The eventual display of these adducts in MHC class II may result in a feed forward loop of further T and B cell activation. Moreover, the accumulation of isoLG adducts within B cells themselves may contribute to their autoreactivity and dysregulation. The accumulation of isoLG adducts within B cells that are positive for the activation marker CD44 suggests a potential functional role of isoLGs in B cell activation and memory. Future studies should focus on the role of isoLG in B cell differentiation and function.

A reduction in anti-dsDNA antibody levels may result from multiple potential mechanisms including a reduction in apoptosis and NET formation (51). The ability of antibodies to bind DNA was discovered in the 1950s in an attempt to define the nuclear antigen inducing anti-nuclear antibody reactivity (52), and anti-DNA positivity is now commonly used to guide therapy in SLE. The specific autoantigen of SLE, however, remains elusive. DNA and nucleosomes are released upon cell death and it has been proposed that, in the presence of other factors including IL-1 and ATP, these act to stimulate an immune response (53). Taken together it is conceivable that scavenging of isoLG reduces apoptosis and thus anti-dsDNA antibody titers. Importantly, bovine serum albumin (BSA) adducted with γ-ketoaldehydes exhibits structural similarity to dsDNA and reacts with the DNA intercalating dyes sybr-green and ethidium bromide(29). Moreover, adducted bovine serum albumin reacts with serum from SLE prone *MRL-lpr* mice in addition to anti-DNA antibodies purified from patients with SLE (29). It is therefore conceivable that isoLG adducted peptides structurally mimic DNA thereby contributing to the production of autoreactive anti-DNA autoantibodies.

SLE is a complex disease associated with hypertension and augmented cardiovascular risk. In the present study, we demonstrate that isoLGs play a causative role in hypertension and autoinflammation in a mouse model of SLE. The importance of isoLGs in the induction of essential hypertension and the associated renal and vascular inflammation suggests a shared pathway of autoimmunity between these conditions. While the role of isoLG in other autoimmune conditions has yet to be studied, scavenging of isoLGs with molecules such as 2-HOBA may be a useful strategy for the treatment of these conditions.

## MATERIALS AND METHODS

### Animals Studied

*C57BL6, NZW, B6.SLE123*, and *NZBWF1/J* were obtained from Jackson’s Laboratory. All experiments were performed on female animals at 32 weeks of age. Acetic acid salt of 2-hydroxybenzylamine (2-HOBA) was synthesized as reported previously (54). Animals were treated with 2-HOBA (1g/L of drinking water) beginning at 7 weeks of age. 2-HOBA was thawed from a frozen stock 3 times per week at which time animal water was changed and was shielded from light in amber water bottles. Radiotelemeters were implanted into animals at 30-weeks of age. Blood pressure was monitored noninvasively using tail cuff and invasively using radiotelemetry as previously described (9, 20). The 4-hour urinary excretion assay was performed utilizing metabolic cages at 30 weeks of age as previously described (7). Animals were weighed and injected with saline at a volume (mL) of 10% of body weight (mg). Urinary volume was measured following 4 hours in the absence of supplemented water. Animals were sacrificed at 32 weeks of age. Urinary albumin and creatinine were measured utilizing commercially available test kits from Exocell. Urinary N-gal was measured using a commercially available ELISA (Abcam). Double-stranded DNA and total IgG were measured using commercially available ELISAs (Alpha Diagonstic Intl).

### Histology

Kidneys were fixed in 10% formalin. A Jones’ sliver stain was performed by the Vanderbilt Translational Pathology Shared Resource. Slides were scanned and evaluated in a blinded analysis. 30-nephrons were scored per kidney for capillary hypercellularity based on the semi-quantitative scale of 0(absent), 1(mild), 2(moderate), and 3(severe). Glomeruli were also scored and the presence of immune complex deposition.

### Flow Cytometry

Tissue homogenates from mice were filtered through a 40 μm filter. Single cell suspensions were stained for flow cytometry and run in three panels using the following antibodies and fluorophores. Pacific LIVE/DEAD Fixable Violet Dead Cell Stain (Thermo Fisher) was used for the lymphoid panel. T cell panel utilized the following surface antibodies: FITC anti-CD45 clone 30-F11 (Biolegend), PE-Cy7 (Cyanain-7) anti-CD3 clone 17A2 (Biolegend), APC (allophycocyanin) anti-CD4 clone RM4-5 (BD Biosciences), Amcyan anti-CD8 clone 53-6.7 (Biolegend). B cell panel utilized the following surface antibodies PE-Cy7 anti-CD45 clone 30-F11 (Biolegend), APC anti-CD19 clone 1D3/CD19 (Biolegend), PE (phycoerythrin) anti-CD44 clone IM7 (Biolegend), Amcyan anti-CD138 clone 281-2 (BD Biosciences). Myeloid panel utilized the Zombie NIR Fixable Viability Kit (Biolegend) and the following surface antibodies: Pacific Orange anti-CD45 clone 30-F11 (Thermo Fisher), BV510 F4/80 anti-F4/80 clone T45-2342 (BD Biosciences), APC anti-MerTK clone 108928 (R&D Systems), AF700 anti-CD11b clone M1/70 (Biolegend), PE/Dazzle 594 anti-CD64 clone X54-5/7.1 (Biolegend), PE anti-Lyc6c clone HK1.4 (Biolegend), PE/Cy7 anti-IAb clone AF6-120.1 (Biolegend), and APC/Cy7 anti-CD11c clone N418 (Biolegend). A known quantity of calibration (counting) beads was added to each sample before analysis. Samples were run on a BD FACSCanto II system or a Cytek Aurora system and analyzed using FloJo software. Gates were set using fluorescence minus one controls. Results were normalized using the bead count and expressed as a number of cells per organ. For isoLG adduct positive cells and plasma cells, cells are represented as total percentage of parent population.

### Intracellular Staining

Following surface antibody staining, cells were fixed and permeabilized using a commercially available cell permeabilization kit (Life Technologies). Intracellular staining for the B cell panel utilized PerCP (peridinin chlorophyll protein) anti-Igκ-Light-Chain clone 187.1 (BD Biosciences). The D11 antibody was labeled with a fluorochrome using the APEX Alexa Fluor 488 Antibody Labeling Kit (Thermo Fisher).

### Anti-IsoLG adduct Antibody ELISA

Whole kidney protein was prepared in non-denaturing lysis buffer (20 mM Tris pH 8.0, 137 mM NaCl, 1.0% NP-40, 2mM EDTA). Un-reacted isoLG was synthesized as previously described (55). 100 μG of protein was incubated with isoLG at a concentration of 100 μM isoLG overnight at 4°C. Immunolon 2HB plates were then coated with D11 at a concentration of 50 μG/mL in coating buffer (1.5 g Na2CO3, 2.93 g NaHCO3, to 1L, pH 9.6) overnight at 4°C at a volume of 100 μL. Plates were then washed x3 with wash buffer (PBS, 0.05% Tween) and then blocked with blocking buffer (PBS + 3% BSA, 3mM EDTA, and 0.1% gelatin) at 37°C for 1 hour. Wells were then incubated with 50 μG/mL isoLG adducted protein in binding buffer ((PBS + 2% BSA, 3mM EDTA, 0.05% Tween-20, 0.1% gelatin) for 1 hour at 37°C at a volume of 100 μL. Plates were then washed x4 with wash buffer. Serum was diluted 1:100 in protein binding buffer and then incubated on wells for 1 hour at 37°C at a volume of 100 μL. Plates were washed x4 with wash buffer and incubated with Protein-G-HRP conjugate (ThermoFisher) at a dilution of 1:100 in secondary antibody diluent (PBS + 1% BSA, 0.05% Tween-20) for 1 hour at 37°C. Plates were washed x3 and 100 μL of TMB substrate solution (ThermoFisher) was added for detection. Plates were incubated at room temperature in the dark for 30-minutes. Stop solution (ThermoFisher) was added and absorbance was measured at 450 nM.

### Studies of isoLG in humans

A cross-sectional study was performed with eligible patients from the VUMC rheumatology clinic. Patients with SLE were recruited as part of a two larger studies. Subjects were recruited as part of the Inflammation Cardiovascular Disease and Autoimmunity in Rheumatic Diseases Study (ICARD). Subjects were 18 years of age or older and met classification criteria for SLE (56). Exclusion criteria include concomitant diabetes mellitus, vaccination against any infectious agent (within 3-months), or active ongoing malignancy. The study was approved by the Vanderbilt Institutional Review Board (IRB # 150544) and all subjects gave written informed consent. Subjects were also recruited as part of the Clearing Lupus Fog with Memantine study. Subjects were between 18-60 years old, met classification criteria for SLE, and scored ≤85 on the RBANS total index (56). Exclusion criteria include recent change in medication (past 4-weeks), regular daily use of opioids/heavy alcohol/other druges of abuse, metabolic derangement (LFT >3x normal or severe renal disease defined as calculated creatinine clearance < 30mL), severe psychiatric disease, refusal to give informed consent, and pregnancy. All subjects gave written informed consent. This study was approved by the Vanderbilt Institutional Review Board (IRB# 180256). Control subjects were recruited by advertisement to the Vanderbilt Clinical Trials Center as part of the Immune Mechanisms of Essential and Lupus-Related Hypertension study. Patients were 18-75 years old. Exclusion criteria included confirmed or suspected renal, renovascular, or endocrine causes of secondary hypertension, concomitant diabetes, concomitant illness requiring corticosteroids or immunosuppressants, recent (within 3-months) vaccination against any infectious agent, active ongoing malignancy, severe psychiatric disorders, or HIV/AIDS. All subjects gave informed consent. This study was approved by the Vanderbilt Institutional Review Board (IRB# 130979). For human PBMC samples, a single cell suspension of PBMCs was prepared and analyzed as previously described (8, 20, 57). PBMCs were harvested by ficoll gradient separation as previously described (16). Live-dead cell staining was performed with 7-aminoactinomycin D. Cells were then stained with the following surface antibodies: PE anti-CD-45 (ebioscience), APC/Cy7 anti-CD14 (BD Biosciences), PE/Cy7 anti-CD11c (ebioscience), APC anti-CD83 (ebioscience), Brilliant Violet 510 anti-CD86 (BD Biosciences). Cells were then permeabilized using a cell permeabilization kit (Life Technologies) and stained with the D11 antibody conjugated to FITC. Conjugation of D11 was performed utilizing the APEX Alexa Fluor 488 Antibody Labeling Kit (Thermo Fisher).

### Mass Spectrometry for isoLG-lysine and superoxide in monocytes

IsoLG-lysine adducts were detected in human monocytes as previously described (58). Superoxide was quantified by monitoring the conversion of dihydroethidium to 2-hydroxyethidium using HPLC as previously described (59).

### Statistics

All data are expressed as mean ± SEM. Comparisons made between 2 variables were performed using Student’s t tests. Normality of distribution of data was confirmed using the D’Agostino-Pearson normality test. Comparisons among more than 2-variables were performed with 1-way ANOVA with Tukey’s post-hoc test. To compare differences in blood pressure 2-way ANOVA followed by post-hoc test was used.

### Study Approval

All animal procedures were approved by Vanderbilt University Medical Center’s Institutional Animal Care and Use Committee, and the mice were housed and cared for in accordance with the Guide for the Care and Use of Laboratory Animals, US Department of Health and Human Services. The institutional review board of Vanderbilt University Medical Center approved the human studies. The subjects signed the written informed consent form before enrolling in the study.

## Data Availability

We will make all data available upon reasonable request.

## Author Contributions

DMP and DGH designed the study. DMP, NV, JPVB, SSD, VNY, SD, AD. LX, AK, and MA performed experiments and acquired the data. DMP, NV, DGH, SD, AD, SSD, and VNY analyzed the data. DMP, MJO, CMS, LJC, and JMW obtained clinical samples and acquired clinical data. ABF provided expertise on renal histology. DMP and DGH wrote the manuscript. All authors reviewed and revised the manuscript. DGH supervised the study.

## Acknowledgments

SOURCES OF FUNDING

This work is supported by National Institute of Health grants 5R35HL140016-02, 5P01HL129941-03, 1R01HL134895-02. Dr. Patrick was funded by the T32 GM007569-44 and is the recipient of a National Institute of Health Ruth L. Kirschstein Individual National Research Service Award (F32HL144050).

